# Pathological neurons generate ripples at the UP-DOWN transition disrupting information transfer

**DOI:** 10.1101/2023.08.01.23293365

**Authors:** Shennan A Weiss, Itzhak Fried, Jerome Engel, Anatol Bragin, Shuang Wang, Michael R. Sperling, Robert K.S. Wong, Yuval Nir, Richard J Staba

**Affiliations:** Dept. of Neurology, State University of New York Downstate, Brooklyn, New York, 11203 USA; Dept. of Physiology and Pharmacology, State University of New York Downstate, Brooklyn, New York, 11203 USA; Dept. of Neurology, New York City Health + Hospitals/Kings County, Brooklyn, NY, USA; Dept. of Neurology, David Geffen School of Medicine at UCLA, Los Angeles, California, 90095, USA; Dept. of Neurosurgery, David Geffen School of Medicine at UCLA, Los Angeles, California, 90095, USA; Dept. of Neurobiology, David Geffen School of Medicine at UCLA, Los Angeles, California, 90095, USA; Dept. of Psychiatry and Biobehavioral Sciences, David Geffen School of Medicine at UCLA, Los Angeles, California, 90095, USA; Brain Research Institute, David Geffen School of Medicine at UCLA, Los Angeles, California, 90095, USA; Depts of Neurology, Epilepsy Center, Second Affiliated Hospital of Medical College, Zhejiang University, Zhejiang, China; Depts. of Neurology and Neuroscience, Thomas Jefferson University, Philadelphia, Pennsylvania, 19107, USA; Department of Physiology and Pharmacology, Sackler School of Medicine, Tel Aviv University, Tel Aviv 6997801, Israel; Sagol School of Neuroscience, Tel Aviv University, Tel Aviv 6997801, Israel; Department of Biomedical Engineering, Faculty of Engineering, Tel Aviv University, Tel Aviv 6997801, Israel; The Sieratzki-Sagol Center for Sleep Medicine, Tel Aviv Sourasky Medical Center, Tel Aviv 6423906, Israel

## Abstract

**Objective:** To confirm and investigate why pathological HFOs (pHFOs), including Ripples [80-200 Hz] and fast ripples [200-600 Hz], are generated during the UP-DOWN transition of the slow wave and if pHFOs interfere with information transmission.

**Methods:** We isolated 217 total units from 175.95 iEEG contact-hours of synchronized macro- and microelectrode recordings from 6 patients. Sleep slow oscillation (0.1-2 Hz) epochs were identified in the iEEG recording. iEEG HFOs that occurred superimposed on the slow wave were transformed to phasors and adjusted by the phase of maximum firing in nearby units (i.e., maximum UP). We tested whether, in the seizure onset zone (SOZ), HFOs and associated action potentials (AP) occur more often at the UP-DOWN transition. We also examined ripple temporal correlations using cross correlograms.

**Results:** At the group level in the SOZ, HFO and HFO-associated AP probability was highest during the UP-DOWN transition of slow wave excitability (p≪0.001). In the non-SOZ, HFO and HFO-associated AP was highest during the DOWN-UP transition (p≪0.001). At the unit level in the SOZ, 15.6% and 20% of units exhibited more robust firing during ripples (Cohen’s d=0.11-0.83) and fast ripples (d=0.36-0.90) at the UP-DOWN transition (p<0.05 f.d.r corrected), respectively. By comparison, also in the SOZ, 6.6% (d=0.14-0.30) and 8.5% (d=0.33-0.41) of units had significantly less firing during ripples and fast ripples at the UP-DOWN transition, respectively. Additional data shows ripple temporal correlations, involving global slow waves, between the hippocampus, entorhinal cortex, and parahippocampal gyrus were reduced by ∼50-80% in the SOZ compared to the non-SOZ (N=3).

**Significance:** The UP-DOWN transition of slow wave excitability facilitates the activation of pathological neurons to generate pHFOs. The pathological neurons and pHFOs disrupt ripple temporal correlations across brain regions that transfer information and may be important in memory consolidation.

**Key Points:**

1. In the SOZ, HFO probability is highest during the UP-DOWN transition of slow wave excitability.
2. In the SOZ, action potentials associated with HFOs occur at the highest probability at the UP-DOWN transition.
3. In the SOZ, a subpopulation of individual units fire more robustly during HFOs at UP-DOWN compared to HFOs at DOWN-UP.
4. Ripple temporal correlations between the hippocampus and other mesial-temporal structures are reduced in the SOZ.

## Introduction

Many patients with focal epilepsy experience more frequent or more severe seizures during non-REM sleep (NREM)^1,2^, and exhibit deficits in memory consolidation^3^. Non-REM sleep is associated with an increase in the rate of inter-ictal epileptiform discharges that may better localize the seizure onset zone (SOZ) ^4,5^. High-frequency oscillations, which are brief (10-100 msec) bursts of spectral power subclassified into ripples (80-200 Hz) and fast ripples (200-600 Hz), also occur more frequently during non-REM sleep^6^. Increased pathological HFO (pHFOs) rates are associated with neuronal injury leading to epileptogenesis^7^, an in particular, increased fast ripple rates can identify brain regions necessary and sufficient for seizure generation^8^, and fast ripples are implicated in seizure genesis^9–11^.

Ripples during NREM sleep are not primarily studied in the context of epilepsy, but rather in the context of facilitating memory consolidation^12–14^. In the hippocampus, ripples during NREM are superimposed on a sharp wave (SpW-R) in local field potential (LFP) recordings. During the SpW-R neuronal ensembles encode experiences from wakefulness that are replayed compressed forward and reversed^15^. Furthermore, ripples important for memory consolidation have also been identified during NREM sleep in association neocortex. These ripples are temporally coupled (*i*.*e*., coincide) with the hippocampal ripples during sleep^13,16–19^ and wakefulness^20^ and promote information transfer and memory. Thus, to understand pathological HFOs (pHFOs) they must first be distinguished from physiological ripples.

During NREM sleep the predominant oscillation is the slow wave^21,22^, which is generated by synchronized membrane potential changes mainly in deep layer pyramidal neurons that repeatedly alternates between a hyperpolarized (DOWN) state to a depolarized (UP) state. The slow wave propagates from the frontal lobe posteriorly, and into mesial temporal structures. Some slow waves can occur globally across the brain, but particularly in mesial-temporal lobe slow waves can be local too^23^. Large membrane potential fluctuations are distinctly lacking in the hippocampus^24^, and it is debated whether hippocampal activity changes there during the slow wave can be classified as DOWN and UP^25^ or DOWN and SpW-R^26^. In murine experiments investigating memory, physiological ripples, in both the hippocampus and neocortex, occur mostly during the DOWN to UP (DOWN-UP) transition^12,13^. In iEEG recordings from patients with epilepsy the relative timing of the UP and DOWN states is inferred by the relative phase of the slow wave, and putatively pathological ripples, in epileptogenic brain regions, are thought to occur relatively more often at the UP to DOWN (UP-DOWN) transition^27-29^. Although physiological and pathological ripples show differences in the preferred angle of coupling to slow wave excitability, there is substantial overlap. Thus, accounting for slow wave-ripple coupling only marginal improves the localization of epileptogenic regions^29-30^. Nevertheless, investigating the neuronal and circuit level mechanisms responsible for pHFOs at the UP-DOWN transition could offer important clues to understanding epileptogenesis and seizure genesis.

We hypothesized that in the SOZ, there are pathological neurons that fire more robustly during pHFOs at the UP-DOWN transition than at the DOWN-UP transition of slow wave excitability. We found, at the group level, that HFO and HFO associated action potentials occur with highest probability at the UP-DOWN transition. However, at the individual unit level in the SOZ, only a minority of the units fired more strongly during HFOs at the UP-DOWN transition. These results confirm that pathologic neurons clusters, embedded in pathological circuits, contribute to generating pHFOs at the UP-DOWN transition of slow wave sleep.

Furthermore, in the SOZ where pHFOs occur, ripple temporal coupling was disrupted between the hippocampus, entorhinal cortex, and parahippocampal gyrus. Since ripple temporal coupling has been associated with information transfer in prior studies^13,16–19^, this observation may indicate a new mechanism contributing to memory disorders in patients with epilepsy.

## Methods

### Dataset collection

All data were acquired with approval from the local institutional review board (IRB) at University of California Los Angeles (UCLA). Six patients with focal epilepsy who were implanted for the purpose of localization of the SOZ in 2009-2010 were included in the study^25,31^. The epileptologist defined SOZ contacts were aggregated across all seizures during the entire depth iEEG evaluation for each patient. Sleep studies were conducted in the epilepsy monitoring unit 48-72 hr after surgery and lasted 7 hr on average, and sleep-wake stages were scored according to established guidelines. The montage included two EOG electrodes; two EMG electrodes scalp electrodes at C3, C4 Pz, and Fz; two earlobe electrodes used for reference; and continuous video monitoring. In each patient, 8–12 depth electrodes were implanted targeting medial brain areas. Both scalp and depth interictal intracranial EEG (iEEG) data from the most medial depth electrode macroelectrode contact, were continuously recorded, during slow wave sleep, with a Stellate amplifier at a sampling rate of 2 kHz, bandpass-filtered between 0.1 and 500 Hz, and re-referenced offline to the mean signal recorded from the earlobes. Each electrode terminated in eight 40-μm platinum-iridium microwires from which extracellular signals were continuously recorded (referenced locally to a ninth noninsulated microwire) at a sampling rate of 28 kHz using a Neuralynx Cheetah amplifier and bandpass-filtered between 1 and 6000 Hz.

### Spike sorting and characterization of high-frequency oscillations HFOs

Action potentials (APs) were detected by high-pass filtering the LFP recordings above 300 Hz and applying a threshold at 5 SD above the median noise level. Detected events were further categorized as noise, single-unit, or multiunit events using superparamagnetic clustering^23^. Unit stability was confirmed by verifying that spike waveforms and inter-spike-interval distributions were consistent and distinct throughout the night. AP time stamps were downsampled to 2000 Hz. HFOs, that were not superimposed on epileptiform spikes, were characterized in the iEEG using previously published methods^32–34^ (Supplementary Methods, https://github.com/shenweiss). HFOs in the microwire LFP recording were not analyzed, reducing the chance that high-frequency events would be influenced by leakage from APs^35^.

### Analysis of the slow-wave, slow-wave associated unit activity and HFOs, and phase standardization

After downsampling the iEEG recordings to 500 Hz. The slow wave was isolated using an optimized Hamming-windowed FIR band-pass filter between 0.1-2 Hz (eegfitnew.m, https://sccn.ucsd.edu/eeglab). In each iEEG channel, we calculated the normalized instantaneous amplitude of the Hilbert transformed band-pass filtered signal and used independent onset and offset normalized minimum amplitude (z-score) and duration criteria, defined from visual inspection of the algorithm results, to identify epochs in which slow oscillatory epochs appeared^29^. We then identified the instantaneous phases of the slow wave, in the proximal iEEG channel, during each HFO and at each AP coinciding with a slow wave epoch. The HFOs were then transformed to a HFO phasor with a single corresponding phase angle and vector length value^29^.

For each unit, we aggregated its APs instantaneous slow wave phase values across the recording duration and tested for circular non-uniformity using Rayleigh’s test (circ_rtest.m) and derived the mean (i.e. preferred) phase angle (circ_mean.m). For each macroelectrode, and corresponding bundle of microelectrodes, we assigned a maximum UP (maxUP) phase angle by identifying the unit, recorded by that bundle, with the largest Rayleigh Z value. That unit’s preferred slow wave angle of maximal AP firing probability was defined as the max UP. We compared the maxUP values across the macroelectrode contacts using a parametric two-way AVOVA for circular data (circ_hktest.m). For all the slow wave HFOs, detected by a single macroelectrode, the maxUP value derived for that macroelectrode trigonometrically adjusted the HFO phasor angles such that θ_HFOadjusted_=θ_maxUP_-θ_HFO_. The instantaneous slow wave phase of the APs, for all the units isolated in the macroelectrode’s corresponding bundle of microelectrodes, were similarly adjusted.

We tested for circular non-uniformity of the HFOs maxUP adjusted phasor angles in the non-SOZ and SOZ and aggregated across macroelectrode contacts and patients, using Rayleigh’s test (circ_rtest.m) and derived the mean (i.e. preferred) phase angle (circ_mean.m). We compared these distributions using the Kuiper’s test (circ_kuipertest.m). This methodology was also used to compare the maxUP adjusted instantaneous slow wave phases of APs coinciding with HFOs.

### Peri-HFO unit activity relative to adjusted slow wave phase angle

All the slow wave HFOs detected in a single macroelectrode contact’s recording were individually compared with the corresponding APs from each individual unit isolated from the LFPs recorded from the bundle of microelectrodes distal to that macroelectrode. APs recorded within the onset and offset of a slow wave HFO was assigned an instantaneous phase of the slow wave that was adjusted by maxUP. Within each individual unit, for each macroelectrode HFO event, a two second raster trial was generated centered at the onset of the HFO event with a resolution of 1 ms (Figure 1,2A). This raster was then convolved with a 100 ms Gaussian kernel and the resulting AP train was down sampled to 40 Hz. These individual gaussian smoothed raster plots were then sorted into 12°,22° bins using the corresponding maxUP adjusted phasor angle of the trial’s HFO event. Within these bins we then calculated the mean gaussian smoothed adjusted peri-HFO firing rate.

**Figure 1:**
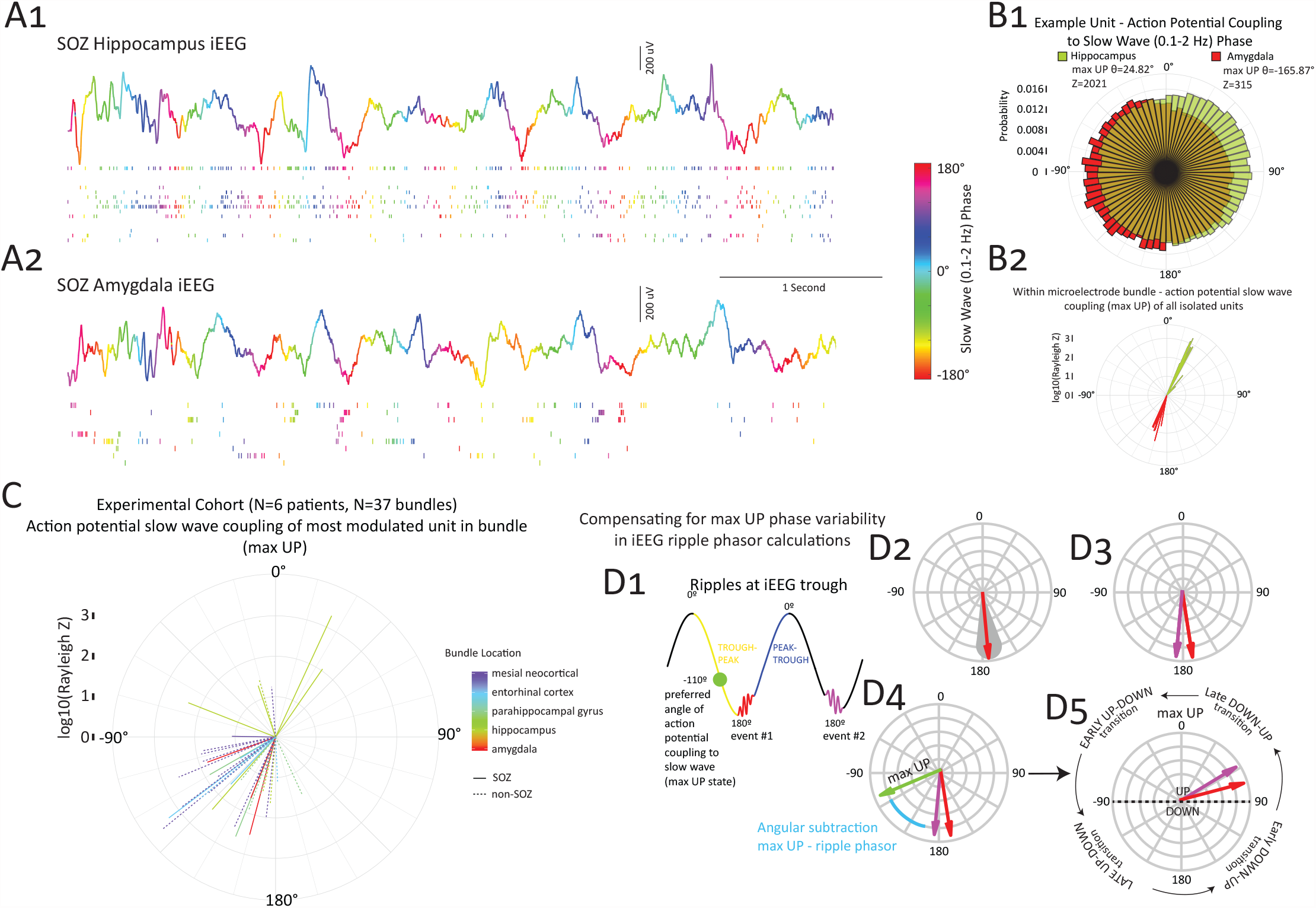
The phase of the maximum of the UP state varies by neuroanatomic location and can be used to adjust and standardize measurements of ripple-slow wave coupling. (A) Simultaneous recordings from Behnke-Fried hybrid macro- and microelectrodes in the hippocampus (A1) and amygdala (A2) that were clinically designated part of the seizure onset zone (SOZ). The iEEG recordings (top) are colored by the instantaneous phase in the slow band (0.1-2 Hz). Note that the peaks are colored red, and the troughs are colored blue, the trough-peak transition is yellow and green, and the peak-trough transition is blue and purple using a circular color scheme. Some local slow waves differ in the two locations. The corresponding raster plots of the units recorded by the microelectrodes are shown (below) and are color coded by the instantaneous phase of the slow wave at time of occurrence. In the hippocampus (A1) the units fire more often from 0-90° (light-dark blue) whereas in the amygdala (A2) the units fire more often from -90° to -180° (red-yellow-green). (B1) Normalized rose plot of all the phase of firing of all action potentials during the entire recording duration of ∼90 min of the most strongly modulated units in A1 (green), and A2 (red) demonstrating the diametrically opposed preferred phase angles of action potential slow wave coupling corresponding to the maximum of the UP state (max UP) and note large Rayleigh Z (Z) scores measuring phase modulation strength. (B2) Polar plot of the max UP phase angle and log10(Z) value for all the units recorded by the Behnke-Fried electrode in (A1) and (A2) showing little variability. (C) Polar plot of the max UP angle and Z value for the unit most strongly modulated by the slow wave isolated from the 37 microelectrode bundles from the Behnke Fried electrodes implanted in six patients. The color of the ray indicates the neuroanatomic location of the microelectrode bundle and solid rays indicate a microelectrode bundle in the SOZ. The neuroanatomic location was significantly correlated with the max UP phase (circular ANOVA, p<0.01), but the SOZ status was not significant (p>0.05). Most neuroanatomic locations, outside the hippocampus, exhibited a max UP from early trough-peak. (D) Illustration of how the max UP phase measured in each bundle can be used to adjust and standardize measurements of ripple-slow wave coupling. (D1) A hypothetical slow wave with two ripple events (red, purple) that occur near the trough, angles shown with the color conventions as in panel A. The green circle indicates the phase of max UP derived from a hypothetical unit, that is set to the trough-peak. (D2) The red ripple can be transformed to a ripple phasor in polar coordinates based on its corresponding slow wave instantaneous phase values. (D3) Polar plot of the two phasors after the transform. (D4) Polar plot of the two ripple phasors with respect to the ray representing that max UP angle. Subtracting the ripple phasor angles from the max UP angle results in an adjusted standardized measurement of the ripple phasor angle with respect to the max UP angle that can be subdivided into stages of transition between actual UP-DOWN and DOWN-UP (D5).

**Figure 2:**
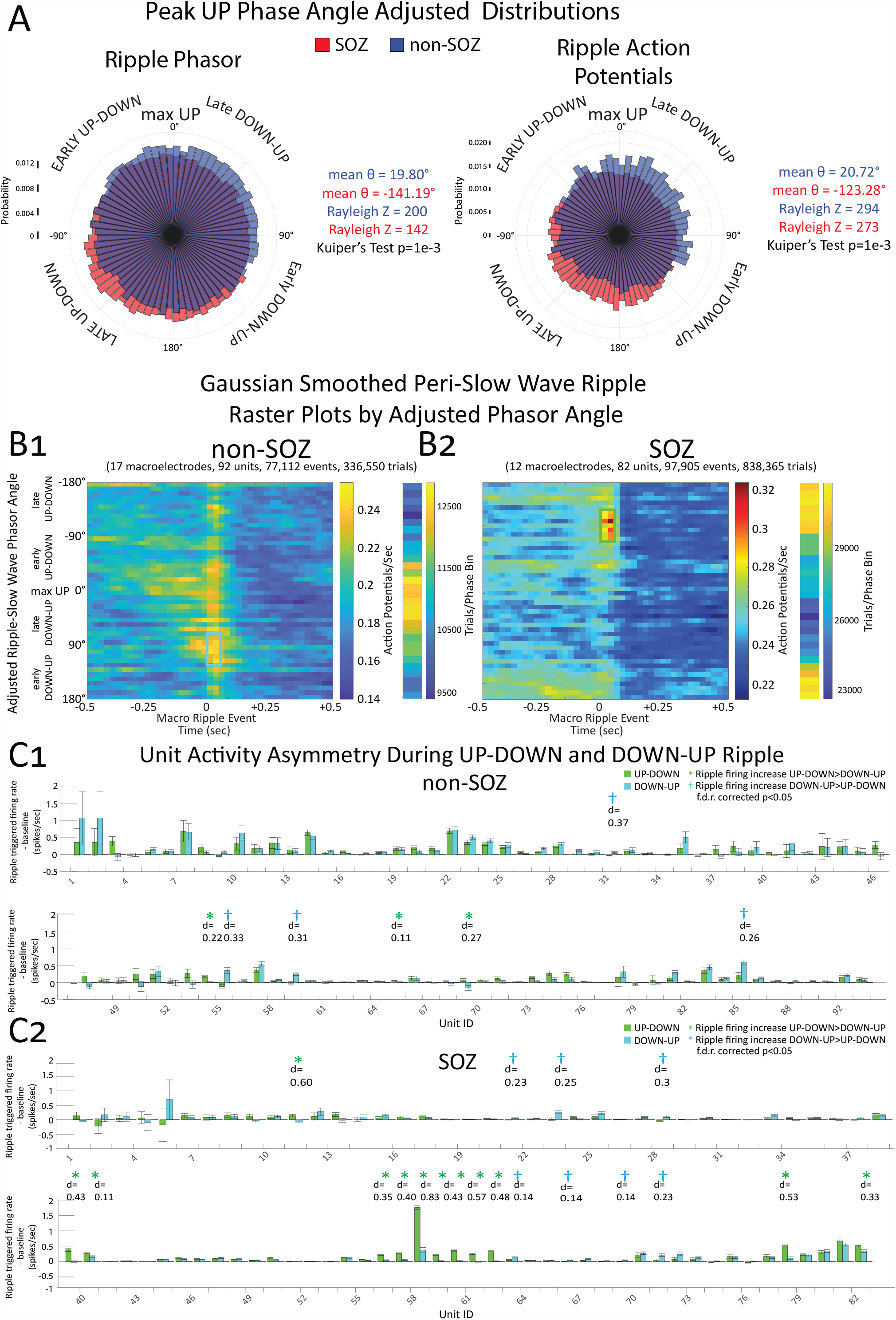
The highest probability of ripples and ripple triggered action potentials (AP), in the seizure onset zone (SOZ) at the group level, is during the late UP-DOWN transition of the slow wave. At the individual neuron level, a minority of SOZ units show significantly enhanced ripple triggered APs at the UP-DOWN transition. (A) Normalized rose plots of the max UP angle adjusted ripple phasors measured during slow wave sleep (left) from electrodes in the SOZ (red) and non-SOZ (blue), also shown are the max UP angle adjusted ripple associated APs instantaneous phase angles with respect to the slow wave (right). The ripple phasors and ripple associated APs in the non-SOZ occur on average during late DOWN-UP, whereas in the SOZ the ripple phasors and ripple associated action potentials occur during late UP-DOWN. (B) Associated, gaussian smoothed peri-slow wave ripple raster plots of units with statistically significant ripple triggered increased firing (p<0.001, f.d.r. corrected) in the non-SOZ (B1) and SOZ (B2). The means of the pooled neuronal firing rate are calculated within 12° bins of the adjusted ripple phasor angle. Mean ripple associated increases in unit firing in the non-SOZ occur at the midway DOWN-UP transition. In the SOZ, ripple associated increases in firing rate are greatest at the late UP-DOWN transition. The number of trials in each phase bin are also shown. (C) For each unit in the non-SOZ (C1) and SOZ (C2), bar plot of the mean ripple triggered firing rate minus baseline firing rate for ripples at DOWN-UP (B1, blue box) and ripples at UP-DOWN (B2, green box). Error bars indicate standard error of the mean (s.e.m). Green asterisks indicate units with significantly (p<0.05, f.d.r. corrected) higher ripple triggered firing during ripples at UP-DOWN compared to ripples at DOWN-UP. Cyan daggers indicate units with significantly higher ripple triggered firing during ripples at DOWN-UP compared to ripples at UP-DOWN.

### Comparison of unit HFO triggered firing during DOWN-UP vs. UP-DOWN

Following gaussian smoothing, for each HFO event-unit trial, the pre-event baseline AP rate was defined as the mean firing rate of the Gaussian smoothed AP train rate beginning 750 msec prior to HFO onset until the event (i.e., bl-fr). The peak HFO-AP rate was defined as the maximum of the Gaussian smoothed AP train rate during the duration of the HFO event (i.e., hfo-fr), and another value hfodiff-fr was defined as hfo-fr minus bl-fr. Within each unit we compared the distribution of hfodiff-fr values for ripples that occurred with an adjusted phasor angle of-1.89 >X>-2.30 radians (UP-DOWN) to the hfodiff-fr values for ripples at 1.05<X<1.47 radians (DOWN-UP) using an unpaired t-test (ttest2.m) with Bonferroni Holms false detection ratio (f.d.r) correction (bhfdr.m) and measured Cohen’s d for effect size (Compute_cohen_d.m). We performed a similar comparison of fast ripples that occurred at -1.57>X>-2.36 radians (UP-DOWN) and -0.4<X<0.4 radians (Max UP).

### Cross-correlogram of ripple timing

We computed the slow wave ripple onset times for all events, or only the events occurring during the UP-DOWN or DOWN-UP transition in single macroelectrode contacts. In select pairs of macroelectrode contacts, the ripple onset times were compared in 1 msec bins using the xcorr.m function and no maximum lag. The cross correlations were normalized to calculate a correlation index by setting the autocorrelations of the sequences at zero lag equal to 1.

## Results

### Patient characteristics and description of data

We analyzed 176 iEEG contact-hours of synchronized macro- and micro-electrode recordings during NREM stage N2/N3 sleep from six patients with medically refractory focal seizures. Of the total patients, 3 were diagnosed with mesial temporal lobe epilepsy (MTLE), one with MTLE and neocortical epilepsy, one with neocortical epilepsy, and one with an unknown site of seizure onset. We detected HFOs in the iEEG, excluding those on epileptiform spikes, and APs from 217 units on the adjacent microelectrodes that occurred on slow waves (Table S1).

### Identification and comparison of maximum UP angle across brain regions

Prior work on slow wave-HFO coupling used the instantaneous phase of the scalp slow wave^27,36^ for comparison to HFO events. This methodology assumes that the slow wave associated changes in excitability during the HFO are global, but slow waves can be locally generated^25^. Prior work has also used the instantaneous phase of the iEEG slow wave^29,30,37^ but this methodology assumes that the slow wave associated changes in excitability during the HFO are uniform irrespective of neuroanatomic location. However, in rat neocortex and entorhinal cortex, due to membrane potential bimodality, maximum pyramidal cell firing occurs during the trough to peak transition of the local slow wave, but in the hippocampus where membrane bimodality is absent, maximum firing can occur during the trough to peak transition or at the peak^24^. Furthermore, in epileptogenic regions, whether neurons maximally fire at other phase angles of the slow wave is unclear.

For reasons noted in the preceding paragraph the first analysis identified for each macroelectrode contact the phase angle of the local iEEG slow wave that was associated with maximum AP normalized probability (Figure 1A,B). This phase was designated as the “max UP” phase (Figure 1B,C). Then the phase angle of the slow wave-associated ripple and fast ripple on the same contact was adjusted to normalize the HFO phase with respect to the max UP state as well as the transitions between UP-DOWN and DOWN-UP (Figure 1D). A similar approach was used to adjust the phase of the APs during the HFOs. The max UP angle was relatively consistent across neuroanatomic locations and occurred during the early trough to peak transition of the slow wave, except in the hippocampus where max UP often was the early peak to trough transition. The max UP angle did not differ for depth electrodes in the SOZ (Figure 1C, two-way circular ANOVA, d.f.=2,8, SOZ X^2^= 3.693, p=0.16, location X^2^=20.327, p=0.01).

### Differences in occurrence of HFOs, and excitability during HFOs, in the SOZ and non-SOZ relative to the UP and DOWN state

We compared the mean angle of slow wave modulation of ripples and ripple-associated APs in the SOZ and non-SOZ. This comparison was also performed after distinguishing ripples in hippocampal and extrahippocampal regions (Figure S1). In all regions, ripples and ripple-associated AP exhibited statistically significant slow wave phase preferences (SOZ, Rayleigh Z=142 [phasor], 273 [AP]; non-SOZ, Z=200,294, p≪0.001 Figure 2A). In the NSOZ, ripples coupled with the slow wave during the late DOWN-UP transition, and maximal ripple-associated AP probability was during middle of DOWN-UP transition (Figs 2A & 2B). By contrast, in the SOZ, ripple coupling and maximal ripple-associated firing probability occurred during late UP-DOWN transition, which was different than the coupling and firing in NSOZ (Kuiper’s V, p=1e-3). These results included regions with interictal epileptiform discharges as part of the non-SOZ, whereas prior investigations included these regions as part of the SOZ^27,30,36^. Thus, in the SOZ, pathological ripples occur during the UP-DOWN transition.

To assess the extent of single units firing during ripples in relation to UP and DOWN phases, we quantified peak firing rate with respect to baseline for ripple-unit trials that occurred at either DOWN-UP phases (Fig. 2B1, blue box) or UP-DOWN phases (Fig 2B2 green box) of slow wave excitability. In the non-SOZ, four units (4.3%, Cohen’s d=0.21-0.27) fired more robustly (t-test, p<0.05, f.d.r. corrected) to ripples at the DOWN-UP transition and three units (3.2%, d=0.26-0.37) fired more robustly to ripples at the UP-DOWN transition (Fig. 2C1). By comparison, in the SOZ, seven units (8.5%, d=0.14-0.30) fired more robustly to ripples at DOWN-UP and thirteen units (15.6%, d=0.15-0.83) fired more robustly to ripples at UP-DOWN (Fig. 2C2).

A similar analysis was performed with fast ripples on slow waves. In the NSOZ, fast ripples coupled with the slow wave during the late DOWN-UP transition and maximal fast ripple-associated unit firing was near max UP (Figs 3A & 3B, p≪0.001). By contrast, in the SOZ, fast ripple coupling (p<0.01) and maximal fast ripple-associated firing (p≪0.001) occurred during late UP-

**Figure 3:**
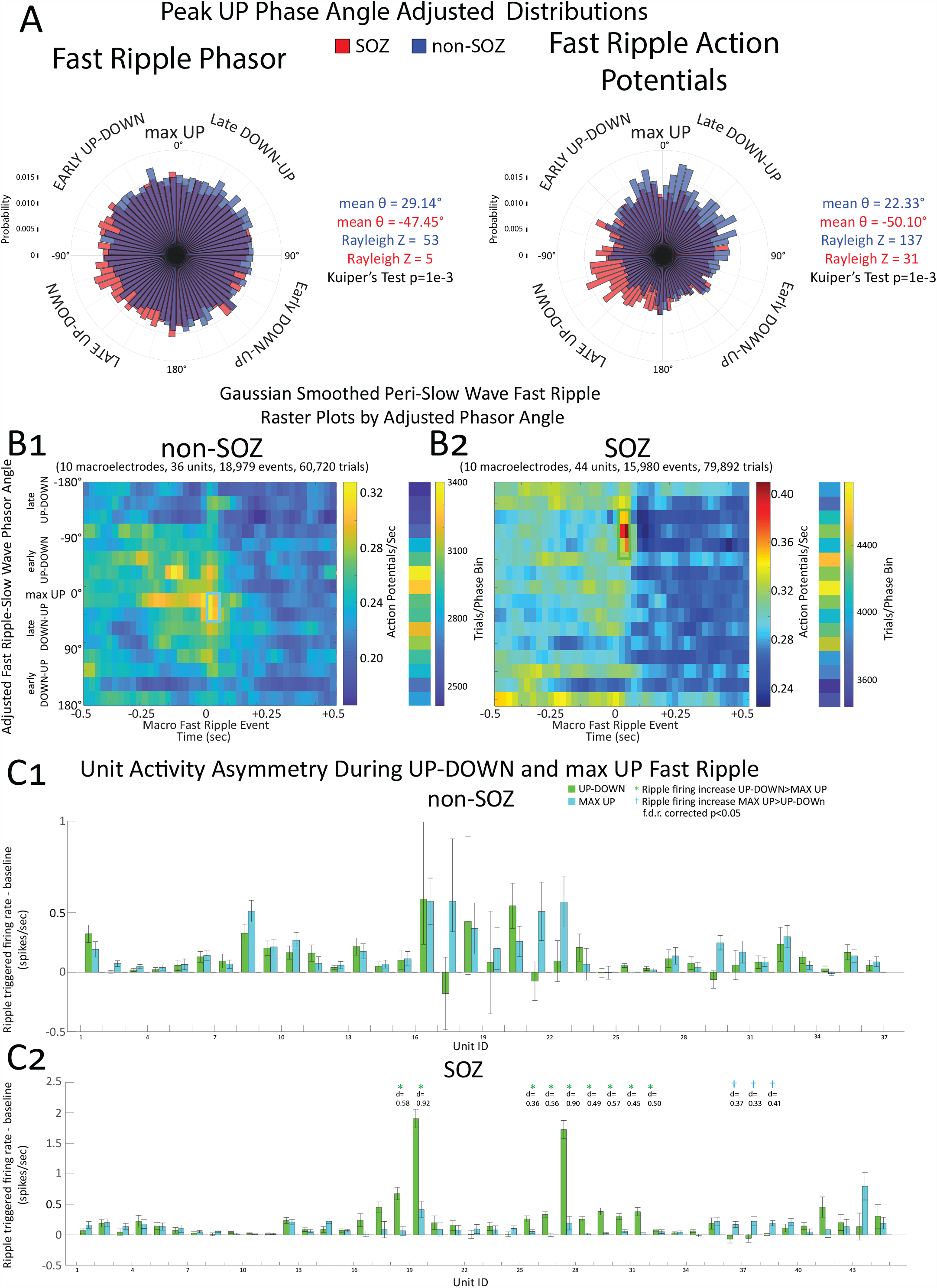
The highest probability of fast ripples and fast ripple triggered action potentials (AP), in the seizure onset zone (SOZ) at the group level, is during the late UP-DOWN transition of the slow wave. At the individual neuron level, a minority of SOZ units show significantly enhanced fast ripple triggered APs at the UP-DOWN transition. (A) Normalized rose plots of the max UP angle adjusted fast ripple phasors measured during slow wave sleep (left) from electrodes in the SOZ (red) and non-SOZ (blue), also shown are the max UP angle adjusted ripple associated APs instantaneous phase angles with respect to the slow wave (right). The fast ripple phasors and fast ripple associated APs in the non-SOZ occur on average during late DOWN-UP, whereas in the SOZ the fast ripple phasors and fast ripple associated action potentials occur during late UP-DOWN. (B) Associated, gaussian smoothed peri-slow wave fast ripple raster plots of units with statistically significant fast ripple triggered increased firing (p<0.001, f.d.r. corrected) in the non-SOZ (B1) and SOZ (B2). The means of the pooled neuronal firing rate are calculated within 22° bins of the adjusted fast ripple phasor angle. Mean fast ripple associated increases in unit firing in the non-SOZ occur near max UP. In the SOZ, ripple associated increases in firing rate are greatest at the late UP-DOWN transition. The number of trials in each phase bin are also shown. (C) For each unit in the non-SOZ (C1) and SOZ (C2), bar plot of the mean fast ripple triggered firing rate minus baseline firing rate for fast ripples near max UP (B1, blue box) and fast ripples at UP-DOWN (B2, green box). Error bars indicate standard error of the mean (s.e.m). Green asterisks indicate units with significantly (p<0.05, f.d.r. corrected) higher fast ripple triggered firing during fast ripples at UP-DOWN compared to fast ripples at near max UP. Cyan daggers indicate units with significantly higher fast ripple triggered firing during fast ripples at near max UP compared to fast ripples at UP-DOWN.

### DOWN transition, which was different than the coupling and firing in non-SOZ (Kuiper’s V, p=1e-3)

With respect to single units that fired more robustly to UP-DOWN fast ripples as compared to max UP fast ripples, in the non-SOZ, no units showed a statistically significant preference (Fig. 3C1). However, in the SOZ three units (6.6%, d=0.33-0.41) showed a preference to max-UP fast ripples, whereas nine units (20.0%, d=0.36-0.90) showed more robust firing to UP-DOWN fast ripples (Fig 3C2).

#### Differences in temporal coupling of ripples in the seizure onset zone

We used normalized cross correlograms to examine the coincidence of ripples between the hippocampus and entorhinal cortex (Figs. 4,S3) or between the hippocampus and parahippocampal gyrus (Fig. S2). In two patients (Figs. 4, S2), we compared the ripple onset cross correlogram of events recorded from contacts in the non-SOZ to that of SOZ contacts. We found that ripple temporal coupling between the hippocampus and entorhinal cortex or hippocampus and parahippocampal gyrus was reduced by 50-80% in the SOZ (Figs 4A, S2A). In another patient with bitemporal epilepsy, we found that ripple temporal coupling was reduced by 80-90%, bilaterally, relative to the non-SOZ as compared to the aforementioned other two patients (Figs. S3A).

**Figure 4:**
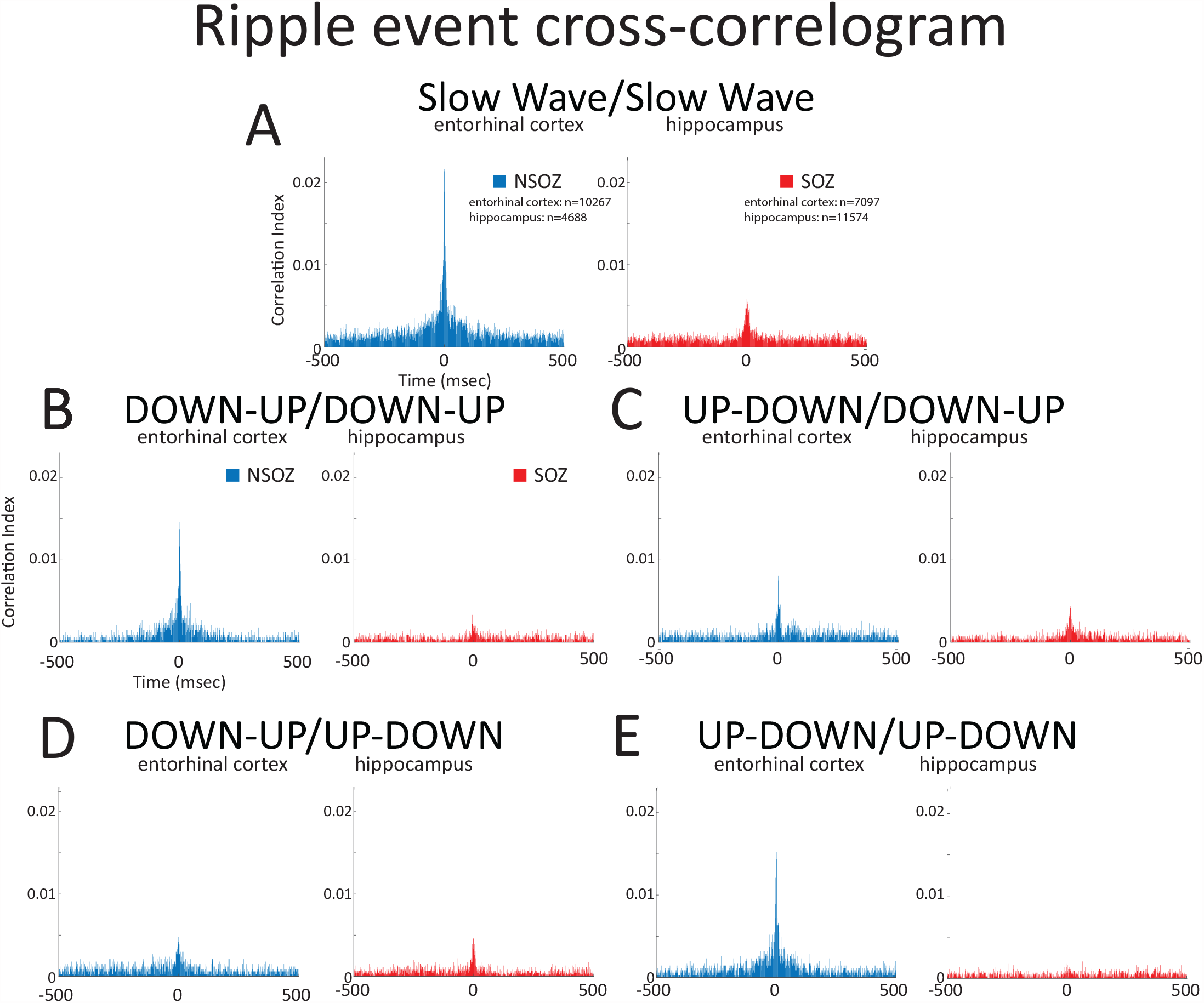
The temporal coupling of ripples during global slow waves is reduced in the seizure onset zone (SOZ). Data from patient 6. Slow wave ripple onset times from the hippocampus and ipsilateral entorhinal cortex were compared using a cross-correlogram. Left sided structures were in the SOZ (red) and right sided structures were in the non-SOZ (blue). (A) Comparison of the normalized cross-correlogram of slow wave ripple onset times between the hippocampus and entorhinal cortex reveals decreased coupling in the SOZ. (B-E) Cross-correlograms calculated after separation of the slow wave ripples by onset times during DOWN-UP transition and UP-DOWN transition. The strongest reduction in ripple coupling in the SOZ, relative to the non-SOZ, was found for ripples that both occurred during the DOWN-UP transition (B), or both occurred during the UP-DOWN transition (E). A smaller reduction was seen in C and D which correspond to ripple temporal coupling involving local slow waves.

After assigning each ripple event to either the DOWN-UP or UP-DOWN transition, in the non-SOZ, we found that ripple temporal coupling was strongest when the events occurred at the same phase of slow wave excitability whether DOWN-UP (Figs 4B,S2B) or UP-DOWN (Figs. 4E,S2E). In the SOZ, ripple temporal coupling was reduced more for these DOWN-UP/ DOWN-UP (Figs 4B,S2B) or UP-DOWN/UP-DOWN pairs (Figs. 4E,S2E) than the temporal coupling of ripples that occurred at opposing phases of slow wave excitability (Fig 4B,E,S2B,E).

## Discussion

To better understand how the UP-DOWN transition of slow wave excitability facilitates pHFO generation, we first established that the phase of the slow wave corresponding to the maximum UP state was similar for units in the SOZ and non-SOZ. Next we showed at the group level in the SOZ, the probability of both HFOs and HFO-associated APs was highest at the UP-DOWN transition. Then, at the single unit level in the SOZ, we found a subpopulation of units that fire more robustly during HFOs at the UP-DOWN transition. Lastly, ripple temporal correlations between the hippocampus and entorhinal cortex, or hippocampus and parahippocampal gyrus showed that ripples coincided less often in the SOZ. These results suggest that the UP-DOWN transition facilitates the activation of pathological neurons to generate pHFOs, and that pHFOs disrupt ripple temporal correlations which may be important in information transfer and memory^13,16–20^.

### Potential Mechanisms Generating pHFOs at the UP/ DOWN transition

A well-supported theory is pHFOs are generated by APs from clusters of pathologically interconnected neurons (PIN)^38–40^. PIN clusters are found following exposure to chemoconvulsants^38,41–43^, but the properties and characteristics that endow PIN cluster neurons to produce pHFOs and seizures are not well understood. In our study, we identified a subpopulation of neurons, primarily in the SOZ, that fired more vigorously during HFOs at the UP-DOWN transition. Most neurons in the SOZ did not show a firing preference for HFOs at different phases of slow wave excitability, and a minority of neurons fired more vigorously during HFOs at the DOWN-UP transition. Thus, the neurons that fire more vigorously during HFOs at the UP-DOWN transition are candidate pathological neurons in PIN clusters because they correspond with the pHFOs at the UP-DOWN transition observed in this and prior studies^27,29,30,36,37^. Notably, our study established that fast ripples occur with a higher probability at the UP-DOWN transition, too. Fast ripples are believed to be pathological irrespective of the phase of slow wave excitability. This complicates the interpretation of the function of the neurons which did not show the firing preference to HFOs at the UP-

DOWN transition and more work is required. By contrast, assigning pathological significance to the neurons in the SOZ that did fire more robustly to HFOs at the UP-DOWN transition^27,29,30,36,37^ is more clear and less ambiguous.

The pathological neurons that fired more during HFOs at the UP-DOWN transition may be facilitated by reduced inhibitory neuron firing^44^ and inhibitory conductances^44,45^ on pyramidal cell soma characterized during the UP-DOWN transition. A decreased inhibitory tone paradoxically shortens the UP state but can, in case of GABA_A_-R blockade, trigger epileptiform discharges at the UP-DOWN transition^46^. An alternative mechanism of pathological HFO generation is that ID2/NKx2.1 DOWN state active neurogliaform inhibitory interneurons may themselves be released from inhibition at the UP-DOWN transition^47^. If pyramidal neurons in epileptogenic tissue exhibit chloride dysregulation and depolarizing inhibitory post-synaptic potentials, as shown previously^11,48–50^, GABA release by these ID2/NKx2.1 at DOWN state onset may contribute to generating pHFOs at the UP/DOWN transition. Experimental epilepsy models are better suited to investigate the role of inhibitory neurons in pHFO generation since patch clamp recordings, microscopy, and optogenetic experiments are likely required^47^.

#### Disrupted ripple temporal coupling due to pHFOs may interfere with memory consolidation

We observed, in the SOZ, ripple coincidence between the hippocampus and entorhinal cortex, or hippocampus and parahippocampal gyrus, was reduced by ∼50-80% as compared to the non-SOZ. Ripple temporal coupling has been investigated in murine models between the hippocampus and association neocortex. Hippocampal SpW-R (i.e., DOWN-UP ripples) induce a DOWN state in association neocortex^51,52^ and the hippocampal SpW-R are also synchronized with neocortical ripples there that occur at the corresponding neocortical DOWN-UP transition^13,19^. This form of ripple temporal coupling has been associated with information transfer and improved memory consolidation^13,51,52^. Ripple temporal coupling has not been investigated within the mesial-temporal lobe in behavioral experiments so the functional implications of reduced ripple coupling in the SOZ is less clear.

In the mesial-temporal non-SOZ, we found substantial ripple temporal correlations for the ripples that occurred either both at the DOWN-UP or UP-DOWN transitions of slow wave excitability. Notably ripple temporal coupling between hippocampus and association neocortex involved only the DOWN-UP ripples, but physiological ripples can be generated in the hippocampus at the UP-DOWN transition too^24^. Weaker coupling was seen for ripples that occurred at opposing phases of slow wave excitability (i.e., DOWN-UP/UP-DOWN).

In the SOZ, relative to the non-SOZ, temporal coupling between ripples at the same phase of slow wave excitability were reduced most. Ripples at opposing phases of slow wave excitability exhibited a smaller reduction in the SOZ relative to the non-SOZ. One explanation is that ripple temporal coupling during global slow waves may be selectively disrupted. More work is needed to understand how the pHFOs in the SOZ mechanistically disrupt the ripple temporal coupling. The pHFOs occur with the highest probability during the UP-DOWN transition but in the SOZ temporal coupling between ripples at the DOWN-UP transition was also reduced.

### Limitations

Comparing the unit activity in the microelectrodes with the slow wave iEEG recorded by the most proximal microelectrode may be inaccurate, because the macroelectrode may be positioned outside the neuroanatomic structure the microwires are recording from, and slow waves may propagate^25^ and exhibit different fields^24^. Analyzing pHFOs in LFP recordings could resolve this issue but introduce AP leakage. Additionally, some local slow wave activity could be inherently pathological^53^. Ripple temporal coupling was analyzed in only three patients because of the requirement of establishing the max UP phase of each macroelectrode contact, and a larger study is required to confirm these findings.

### Conclusion

In epileptogenic regions, the UP-DOWN transition of slow wave excitability facilitates the activation of pathological neurons to generate pHFOs. We found that these pathological neurons represent only 15-20% of the units sampled in the SOZ. Such pathological neurons may be excited at the UP-DOWN transition by decreased inhibitory tone or possibly depolarizing inhibition. Epileptogenic mesial-temporal regions also exhibit decreased physiological and pathological ripple coincidences. Ripple temporal correlations between brain areas may enable information transfer and memory consolidation. Decreased ripple temporal correlations within epileptogenic regions, and across structures, may be due to pathological neurons that abnormally generate pHFOs at the UP-DOWN transition or the pHFOs themselves. Decreased ripple temporal correlations may be an important mechanism in memory impairment in patients with epilepsy.

## Acknowledgements

We thank Dr. Gyorgy Buzsaki, Dr. Anli Liu, and Dr. Simon Henin for their helpful suggestions prior to drafting the manuscript. Epilepsia ethical publishing statement: “We confirm that we have read the Journal’s position on issues involved in ethical publication and affirm that this report is consistent with those guidelines.”

## Data availability statements

The processed data used to generate the figures and statistics in this paper are available at https://zenodo.org/record/8127018, the code is available at https://github.com/shenweiss/publishedcode/tree/master/UPDOWNHFO. The raw data is available upon reasonable request to Dr. Richard Staba or Dr. Yuval Nir.

## Contributions

Shennan A. Weiss: Conceptualization, Methodology, Software, Investigation, Resources, Writing – original draft, Writing – review & editing, Funding acquisition. Itzhak Fried: Resources. Jerome Engel: Writing – review & editing, Funding acquisition. Shuang Wang: review & editing. Michael R. Sperling: Writing – review & editing, Funding acquisition. Yuval Nir: Conceptualization, Methodology, Software, Investigation, Resources, Writing – review & editing. Richard J. Staba: Conceptualization, Methodology, Writing – review & editing, Funding acquisition.

