## supplementary data for "Pathological neurons generate ripples at the UP-DOWN transition disrupting information transfer"

### Supplementary Methods

*HFO Detection*

A two-stage algorithm first used a custom Hilbert based detector implemented in Matlab with artifact rejection features to identify transient elevations in ripple (80-200 Hz) and fast ripple (200-600 Hz) amplitude. In the second stage, the different HFO types: (1) ripples on oscillations (RonO); (2) ripples on spikes (RonS); (3) fast ripples on oscillations (fRonO); (4) fast ripples on spikes (fRonS), and (5) sharp-spikes without a HFO were distinguished using the topographical analysis of the wavelet convolution[22,34,36]. In brief, this method identifies contours of power in the wavelet time-frequency transforms. Contours corresponding to power values less than a threshold defined by 0.2 * ($\text{max}_{\text{time-frequency}}$-power – $\text{min}_{\text{time-frequency}}$-power) + $\text{min}_{\text{time-frequency}}$-power were removed. Each of the remaining contours was subsequently classified as a closed loop contour (CLC) if the contour's first and last vertex coordinate was identical, and an open loop contour (OLC) if the first and last vertex were distinct. HFOs on spikes were distinguished by comparing onset of the outermost closed-loop isopower contour of the HFO in the time-frequency spectrogram with onset of the outermost open-loop isopower contour of the spike and determining if the latter occurred earlier. Open-loop contours without closed loop contours were designated sharp-spikes without HFOs. This method also identified the spectral content, power, and duration of each HFO. Ripples and fast ripples were distinguished by the mean frequency of the closed-loop contour group, and peak spectral content and power from the largest isopower contour value in the closed-loop contour group. Duration was computed from the onset and offset of the outermost contour in the closed-loop contour group. Compared to visual analysis of two reviewers, HFO and spike detection sensitivity and accuracy of event detection, using this method, was typically 80–90%. Following automatic detection of HFO and sharp-spikes, false detections of clear muscle and mechanical artifact were deleted by visual review in Micromed Brainquick. HFOs on spikes were excluded from the analysis.

### Supplementary Tables

Table S1: Clinical demographics and a full description of the brain structures and number of units identified in the recordings. Abbreviations: L, left hemisphere; R, right hemisphere; H, hippocampus; A, amygdala; EC, entorhinal cortex; PHG, parahippocampal gyrus; TG, temporal gyrus; AC, anterior cingulate; MC, middle cingulate; PC, posterior cingulate; OF, orbitofrontal and medial prefrontal cortex; SM, supplementary motor area; P, parietal cortex; TO, temporo-occipital, PT, posterior temporal cortex; FG, fusiform gyrus; IFG, inferior frontal gyrus

| Patient/ Age/ Gender | PET/MRI Findings | Seizure onset zone (SOZ) | Resection/ Outcome | Macroelectrode/units |
|---|---|---|---|---|
| 1. | Metabolic and structural abnormalities left temporal. Pathology FCD IB | Left temporal spreading to right entorhinal cortex | Left temporal, significant improvement | L H :, L A :, L PHG : 6 units, L AC : 9 units, L TO :, R H : 5 units, R A : ,R EC : 11 units, R PHG : 2 units, R AC :, R TO : |
| 2. | Mild metabolic abnormality left temporal, structural abnormality right frontal lobe and left hippocampal T2 hyperintensity | Left medial temporal lobe | No surgery | L H : 6 units, L A : 8 units, L PHG : 13 units, L AC : 3 units, L SMA, R H : 8 units, R A : 8 units, R PH : 6 units, R AC : 4 units, R OF, R SMA |
| 3. | Mild metabolic abnormality right temporal, mild structural abnormality left temporal | Bilateral mesial temporal lobe and right inferior frontal gyrus | No surgery | L H : 3 units, L A :, L EC : 1 unit, L AC : 3 units, L OF : 10 units, R H : 16 units, R A : 8 units, R EC : 7 units, R TG : 12 units, R AC :, R OF :, R IFG : |
| 4. | Metabolic abnormality right parietal lobe, normal structural. Pathology FCD IIA & IIB | Right parietal | Right parietal seizure free | L H : , L A :, L PHG : 4 units, L P: 8 units, R H: 9 units, R A:, R PH: 8 units, R P: 7 units |
| 5. | Volume loss due to prior LAMTL. Normal around margins of resection site and elsewhere | Not localized | No surgery | L PH : 8 units, L AC : 1 unit, L OF : 11 units, L SMA : 8 units, L PT : 2 units, R H : 5 units, R PH : 6 units, R OF : 7 units, R P : 10 units |
| 6. | Metabolic and structural abnormalities left temporal including amygdala T2 hyperintensity. Pathology gliosis | Left mesial temporal lobe | Left temporal seizure free | L H : 5 units, L A : L EC : 8 units, L OF : 7 units, R H : 1 units, R A :, R EC : 11 units, R OF : |

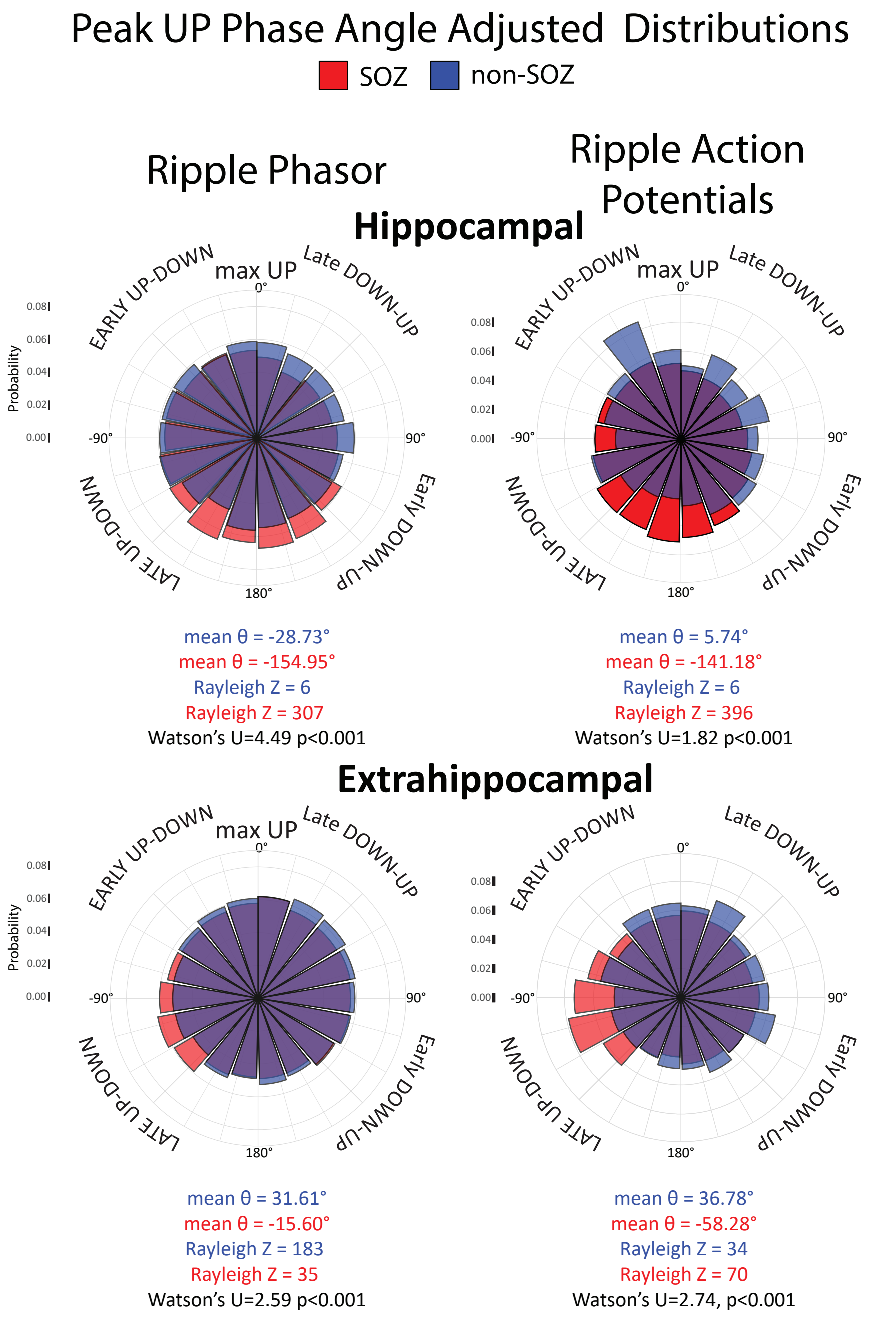


Figure S1: Distinguishing recording sites in hippocampus and extrahippocampal structures demonstrates that pathological ripples in the hippocampus also occur during the UP-DOWN transition. Normalized rose plots of the max UP angle adjusted fast ripple phasors measured during slow wave sleep (left) from electrodes in the SOZ (red) and non-SOZ (blue), also shown are the max UP angle adjusted fast ripple associated action potential instantaneous phase angles with respect to the slow wave (right) measured from recording sites in the hippocampus (top) and extrahippocampus (bottom). Note also that the mean phase angle of action potential firing in the hippocampal non-SOZ is near max UP, whereas in extrahippocampal structures it is late DOWN-UP. In the SOZ, mean action potential firing occurs at late UP-DOWN in the hippocampus but early UP-DOWN in the neocortex. Watson's U test was performed using the circular package in R.

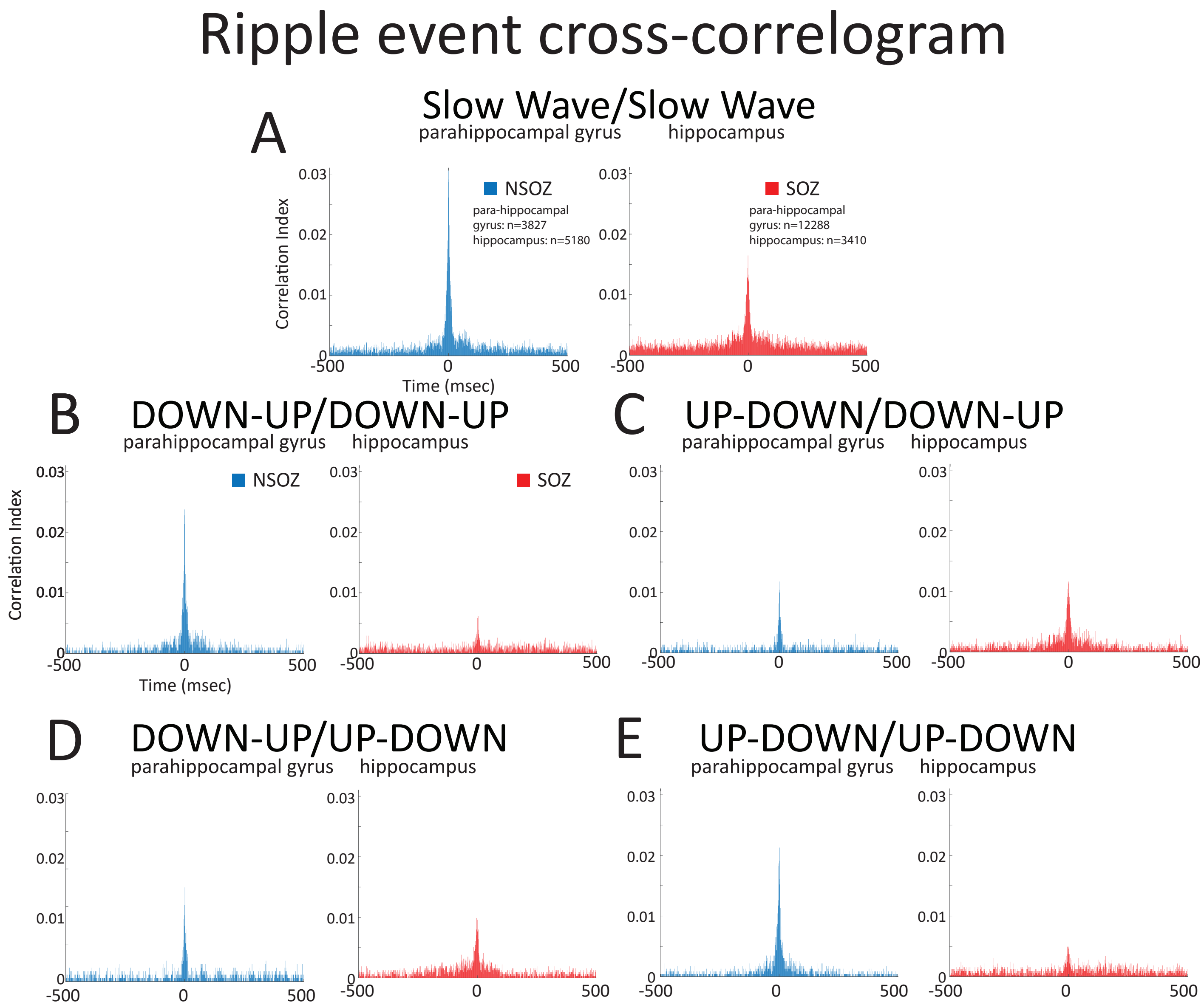


Figure S2: The temporal coupling of ripples is reduced in the seizure onset zone (SOZ) due to a reduction in ripples that couple together at the DOWN-UP and UP-DOWN transition. Data from patient 2. Slow wave ripple onset times from the hippocampus and ipsilateral parahippocampal gyrus were compared using a cross-correlogram. Left sided structures were in the SOZ (red) and right sided structures were in the non-SOZ (blue). (A) Comparison of the normalized cross-correlogram of slow wave ripple onset times between the hippocampus and parahippocampal gyrus reveals decreased coupling in the SOZ. (B-E) Cross-correlograms calculated after separation of the slow wave ripples by onset times during DOWN-UP transition and UP-DOWN transition. The strongest reduction in ripple coupling in the SOZ, relative to the non-SOZ, was found for ripples that both occurred during the DOWN-UP transition (B), or both occurred during the UP-DOWN transition (E).

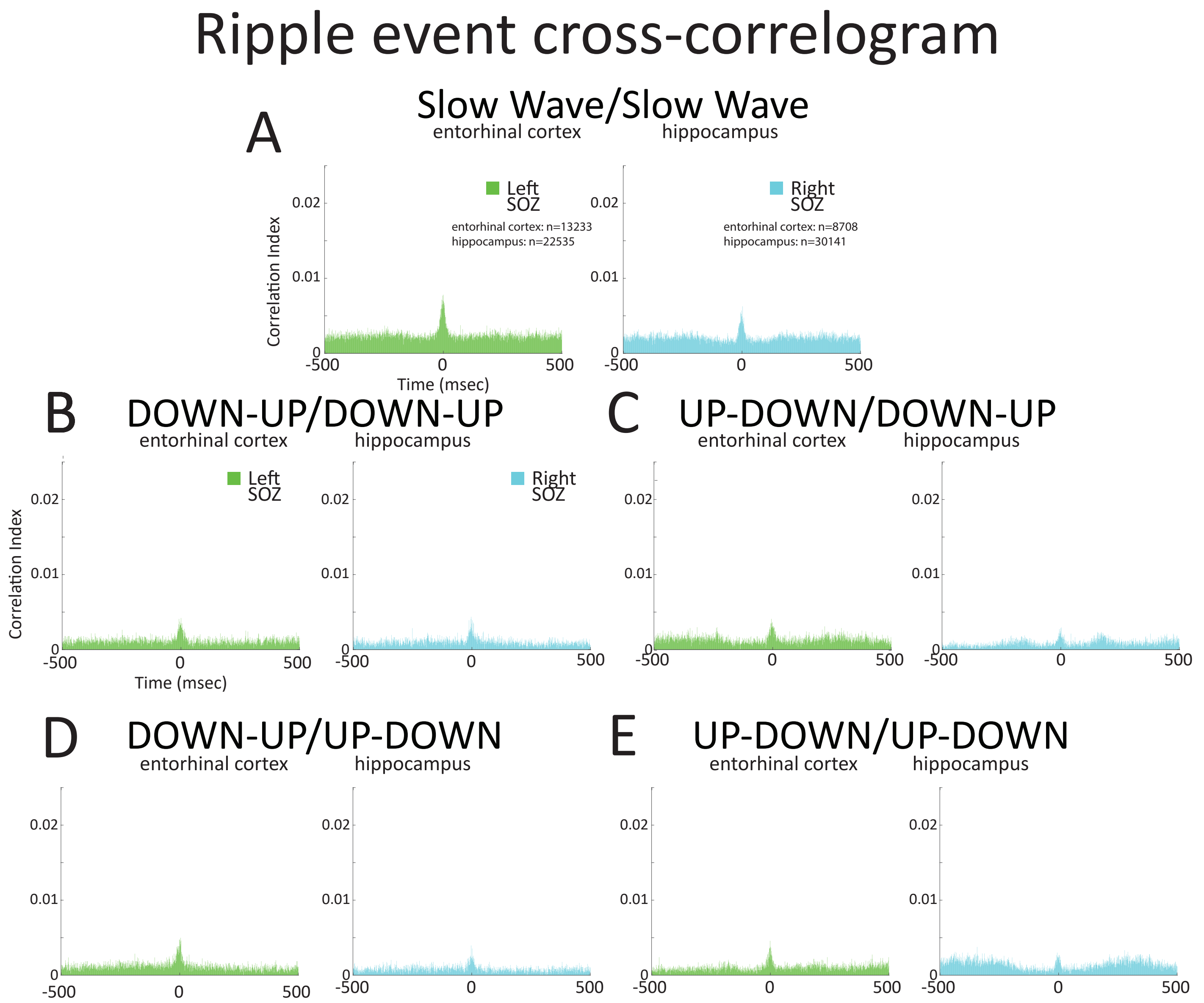


Figure S3: The temporal coupling of ripples is reduced in the seizure onset zone (SOZ) due to a reduction in ripples that couple together at the DOWN-UP and UP-DOWN transition. Data from patient 3. Slow wave ripple onset times from the hippocampus and ipsilateral entorhinal cortex were compared using a cross-correlogram. Left sided structures were in the SOZ (green) and right sided structures were also in the non-SOZ (cyan). (A) Comparison of the normalized cross-correlogram of slow wave ripple onset times between the hippocampus and entorhinal cortex reveals decreased coupling in the two SOZ relative to the non-SOZ shown in Figure 7A in blue. (B-E) Cross-correlograms calculated after separation of the slow wave ripples by onset times during DOWN-UP transition and UP-DOWN transition.
